# Explanatory factors for the occurrence of the cholera epidemic in the Kadutu health zone, city of Bukavu in the Democratic Republic of Congo

**DOI:** 10.1101/2025.01.20.25320821

**Authors:** Bonhomme Kalimira Kachelewa, Denise Ngondo, Henry Mata, Jack Kokolomami, Thiery Lengu Bobanga, Jean Nyandwe Kyloka

## Abstract

**Background:** The Kadutu Health Zone (HZK) in Bukavu, located in the east of the Democratic Republic of Congo, is one of the areas most affected by cholera. This study aims to identify the factors explaining the occurrence of cholera epidemics in this region.

**Methods:** A case-control study was conducted between January and June 2024, with a sample drawn from cholera cases of the 2023 outbreak. A questionnaire was administered to 112 cases and 224 controls. Data were analyzed using SPSS v 25 software.

**Results:** The results revealed nine factors significantly associated with the onset of the epidemic: low monthly income (OR = 2.49; p = 0.010), non-washing of hands before eating (OR = 25.85; p < 0.001) and before breastfeeding (OR = 3.06; p = 0.006), lack of hygiene of soiled hands (OR = 19.37; p < 0.001), defecation outside the toilet (OR = 4.54; p < 0.001), recourse to traditional practitioners (OR = 3.28; p = 0.033), lack of toilets (OR = 2.07; p = 0.017), and lack of knowledge of the modes of transmission (OR = 2.94; p = 0.032) and prevention (OR = 1.75; p = 0.037).

**Conclusion:** Cholera prevention in the HZK requires a multisectoral approach, taking into account sociodemographic, economic, environmental, cultural and health factors.

## I. INTRODUCTION

Cholera is a strictly human contagious diarrheal disease. It is caused by a Gram-negative bacillus, *Vibrio cholerae*, of serogroups O1 and O139. This disease of fecal peril par excellence is a real public health emergency [1,2]. It is a disease of dirty hands attributed to the poor. Cholera is a favorite companion of natural disasters and conflict situations with mass population displacement [3]. Worldwide, in 2022, 80 countries had reported data on cholera to the World Health Organization (WHO). Among them, 44 countries had reported a total of 472,697 cases and 2,349 deaths, representing a case fatality rate of 0.5%. [4]. Since 2007, the Democratic Republic of Congo (DRC) has launched a vast program to combat cholera. The aim of this program was to eliminate cholera in the country by 2030. The threshold for this elimination will be one (confirmed) case per 100,000 inhabitants on a national scale [5].

The DRC has been plunged into instability for several years due to incessant wars and conflicts. This situation is more pronounced in the eastern part of the republic. These events negatively impact several sectors of economic life on a national scale [5]. Lack of access to drinking water, sanitation and hygiene (WASH). Poverty, conflicts, population displacement, natural disasters, climate change, insufficient latrines, human-to-human contact by asymptomatic and symptomatic carriers are very recurrent in the displaced persons camps and in the Kadutu health zone (HZ). Our humble conviction is that the combination of the factors listed above may be the pivot of the resurgence of incessant cholera epidemics.

Since the beginning of 2023 until November 19, 2023, the province of South Kivu has reported 8,374 cases of cholera, including 103 deaths, i.e. a lethality of 1.25%. In the city of Bukavu, the HZ of Kadutu is in the lead with 855 cases including 9 deaths, i.e. a lethality of 1.05%. The HZ of Ibanda itself had a cumulative total of 529 cases including 5 deaths, i.e. a lethality rate of 0.94% and the HZ of Bagira 279 cases including 1 death, i.e. a lethality of 0.34% [6].

The choice of this theme is motivated by the increasing importance that the fight against cholera has in the national health policy nowadays. Although appropriate control measures have been taken, the occurrence of cholera in the Kadutu HZ requires a new approach adapted to its endemic-epidemic nature. This nature is motivated by the lack of previous studies and the need for reliable local data.

## II. PATIENTS AND METHODS

### Type and period of the study

This is a case-control study which was conducted to determine the explanatory factors for the occurrence of the cholera epidemic in the Kadutu Health Zone during the period from January to June 2024.

### Study setting

The study focused on the Kadutu HZ in the city of Bukavu, which became autonomous in 2003 after the division of the former urban HZ of Bukavu. It covers 15 km² with a population of 429,505 inhabitants spread across 13 health areas (AS) with a density of 28,634 inhabitants/Km ^2^ [7].

### Study population

The study population consisted of cholera patients (cases) from the May to July 2023 epidemic who were on the linear cholera lists. Non-sick subjects (controls) are people with the same characteristics except for the disease as the sick subjects.

### Sampling

The sample size was determined using the Statcalc function of Epi Info software version 7. The proportion of exposed subjects among controls was estimated at 50% (Kashinde M. et al. 2023) [8].

Confidence interval at the 95% confidence threshold (95% CI) was chosen by default by the computer. This implies that the significance threshold is 5%. The power of the test is 80%, which implies that the ꞵ is 20%. The ratio is 1 case for 2 controls. The Odds ratio = 2 (Kashinde M. et al. 2023) [8].

With all these values introduced into the statcal function of Epi Info 7, the sample size for the present study is 335 subjects. By adding 10% to compensate for data entry errors and outlier responses, the sample size will be increased to 370 subjects. All things considered, we will therefore have 125 cases and 245 controls. During data collection, 336 people were surveyed, divided into 112 cases and 224 controls.

### Data collection

Cases were selected from linear lists, after excluding irrelevant criteria. A random draw was made on this basis, with a sampling step determined by the sample size. Controls were chosen from households or direct neighbors of the cases, and, in the absence of a control, from the neighboring plot. Each case was matched with two controls.

### Variables studied

The dependent variable is the cholera epidemic. The independent variables are grouped into four categories: sociodemographic (age, sex, marital status, residence/displaced, promiscuity), socioeconomic (occupation, religion, household size, education level, income), environmental (water situation, sanitation, hygiene, seasons, toilet floor) and cultural/health (traditional medicine, handling of corpses, defecation, knowledge of cholera, surveillance, vaccination).

### Statistical analysis of data

Data collected via Kobo Collected data were exported to Excel format, then processed and analyzed with SPSS for Windows 25. Qualitative and quantitative variables were analyzed in terms of frequencies, percentages, central tendencies and dispersion. Bivariate analysis used Pearson’s Chi-square test to identify significant associations with cholera. Odds ratio (OR) and its 95% confidence interval (95% CI) at the threshold of p < 0.05 were used to assess measures of association and impact. Logistic regression was used to adjust independent variables and determine factors significantly associated with the occurrence of cholera epidemic in Kadutu HZ.

### Ethical Consideration

The research protocol was submitted to the ethics committee of the School of Public Health of Kinshasa (ESPK) and received its approval under No. ESP/CE/189/2023. The research was conducted in strict compliance with the main ethical principles which are: respect for the person (autonomy and self-determination), and benevolence in justice.

## III. RESULTS

### III.1. Socio-demographic characteristics of respondents in the Kadutu HZ

The results shown in this figure1 indicate that the majority of respondents, 84 or 25%, reside in the Maria/Karhale Health Area(HA), followed by the Ciriri 2 HA with 69 subjects, representing 21%. The Biname HA had only 3 respondents, accounting for 1%.

**Figure 1:**
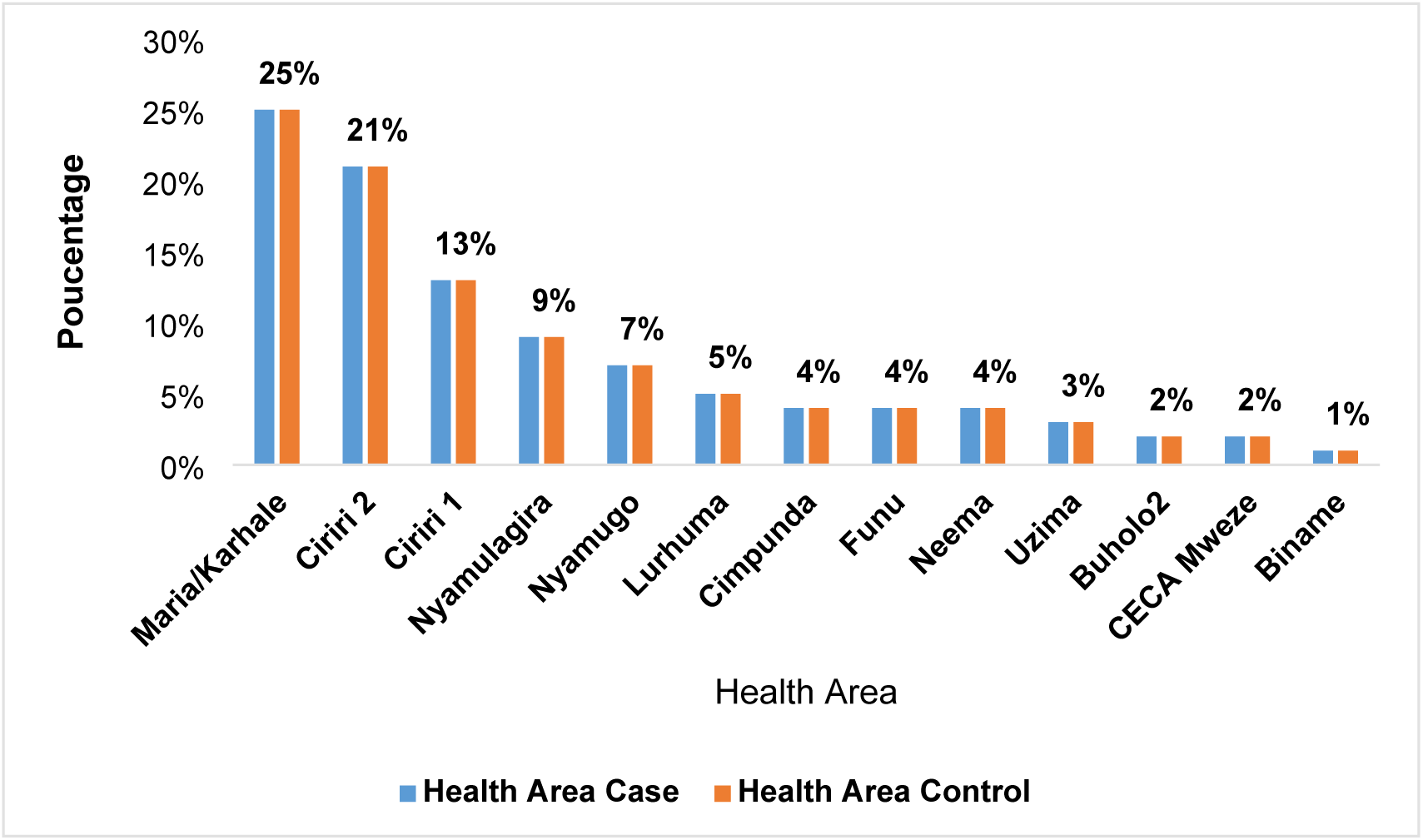
Distribution of cases and controls by health areas in the Kadutu health zone in the city of Bukavu.

### III.2. Health Zone Sociodemographic and economic characteristics of respondents

The mean age of cases and controls was 29 years ±11.5 and 26 years ±10.6 respectively. Table I shows that all age groups were recruited for the survey. But the group that contains many candidates is that of 25 to 34 years, that is to say a proportion of 32% of cases and 28% of controls. The female sex represents 54% of cases and 53% of controls. Married people represented 47% of cases and 42% of controls, followed by single people 46% of cases and 54% of controls. As for the profession, 25% of cases and 28% of witnesses do small businesses followed by the unemployed 21% of cases as well as witnesses. Housewives represent 19% of cases and 14% of witnesses. The Catholic religion accounts for 54% of cases and 50% of witnesses. Revival churches account for only 7% of cases and 3% of witnesses.

**Table 1:**
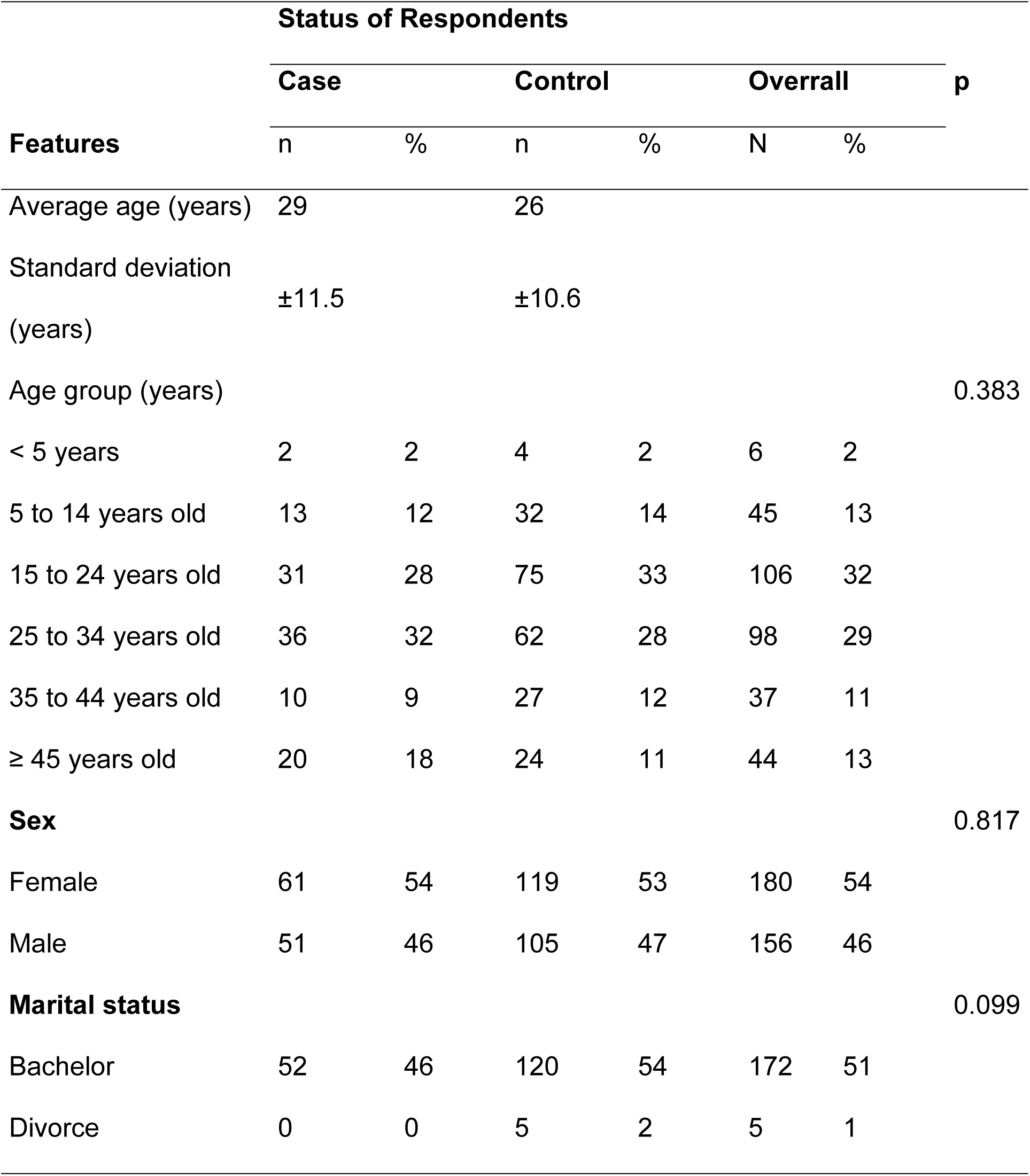
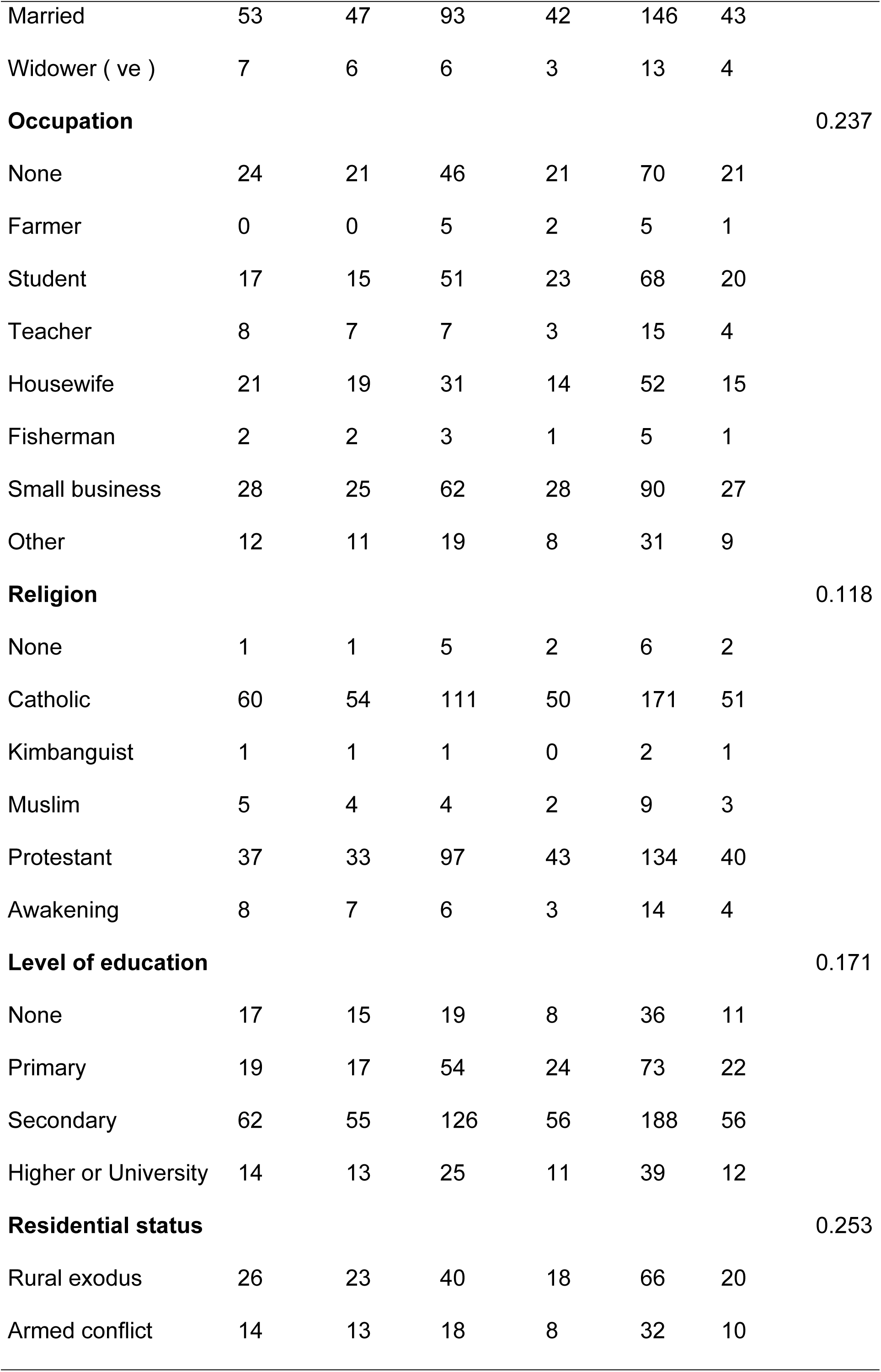
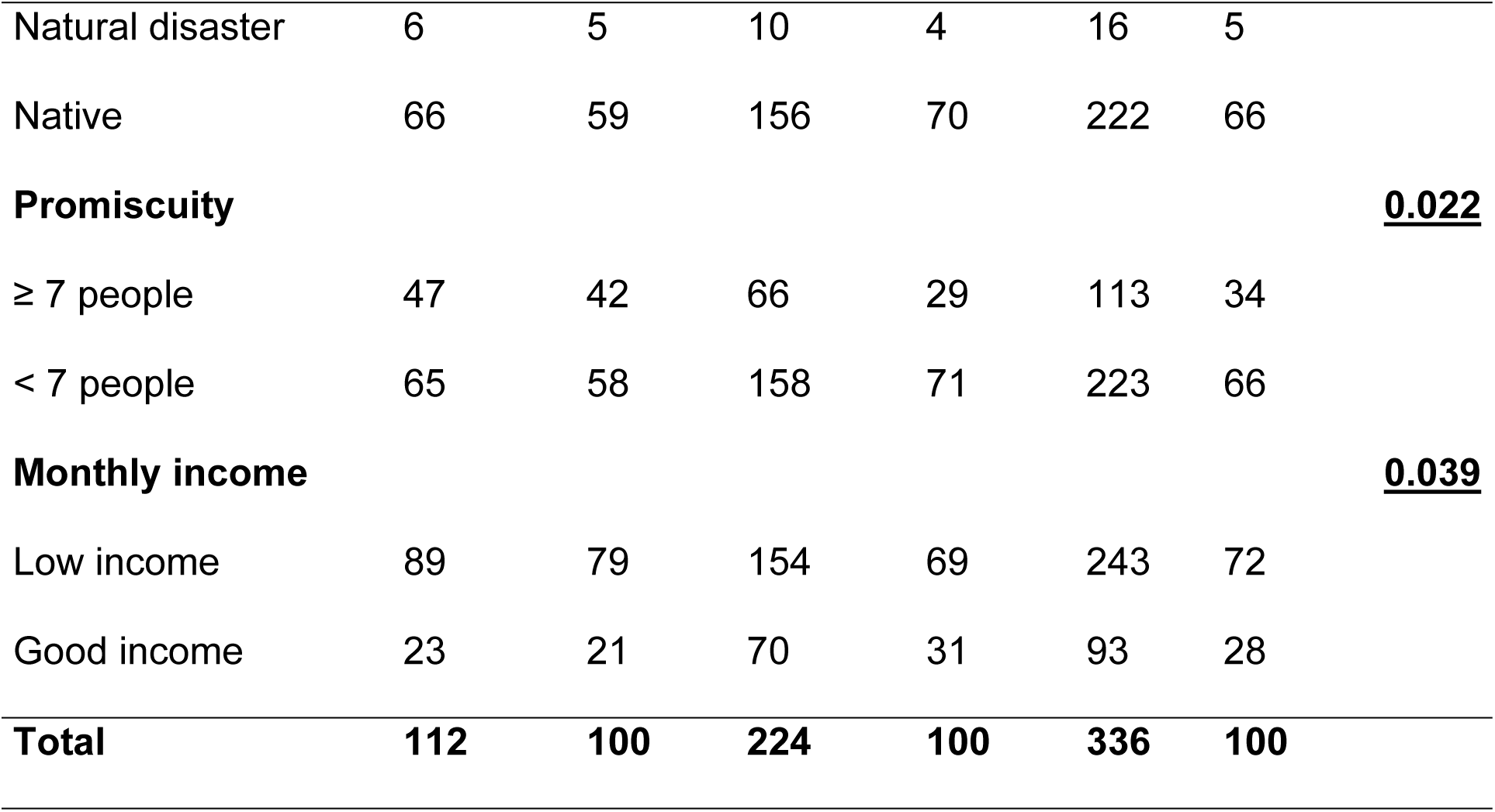
Sociodemographic and economic characteristics of respondents.

The secondary education level includes 55% of cases and 56% of controls. The uneducated represent 15% of cases and 8% of controls.

More of the respondents in the HZ are indigenous, i.e. 59% of cases and 70% of witnesses. Those displaced by natural disasters, armed conflicts or rural exodus represent 41% of cases and 30% of witnesses.

Household promiscuity is known by 58% of cases and 71% of controls. Promiscuity and low income are significantly associated with the cholera outbreak in the Kadutu HZ

### III.3. Environmental factors of respondents in the Kadutu HZ

Table 2 shows that 30% of cases and 24% of controls obtain their drinking water from an unprotected source. While 23% of cases and 22% of controls obtain their drinking water from a public tap/terminal and/or fountain. As for obtaining water from a tap in the neighboring plot, 19% of cases and 26% of controls obtain water from a tap in the neighboring plot. On the other hand, 15% of cases have taps in their plot and 26% of controls. Only 2% of cases use rainwater or surface water as do 1% of controls. The place where water is obtained is not associated with the occurrence of cholera in the Kadutu HZ among respondents (p = 0.636). Forty percent of households where there is a cholera patient use unclosed containers compared to 79% of controls. This practice is associated with the cholera epidemic (OR = 2.556 [1.525-4.282] and p < 0.001); Regarding the treatment of drinking water, 58% of cases do not treat drinking water compared to 62% of controls. Water treatment is not a factor significantly associated with the cholera epidemic in the Kadutu HZ (OR = 0.861; 95% CI [0.543-1.367], p = 0.528).

**Table 2:**
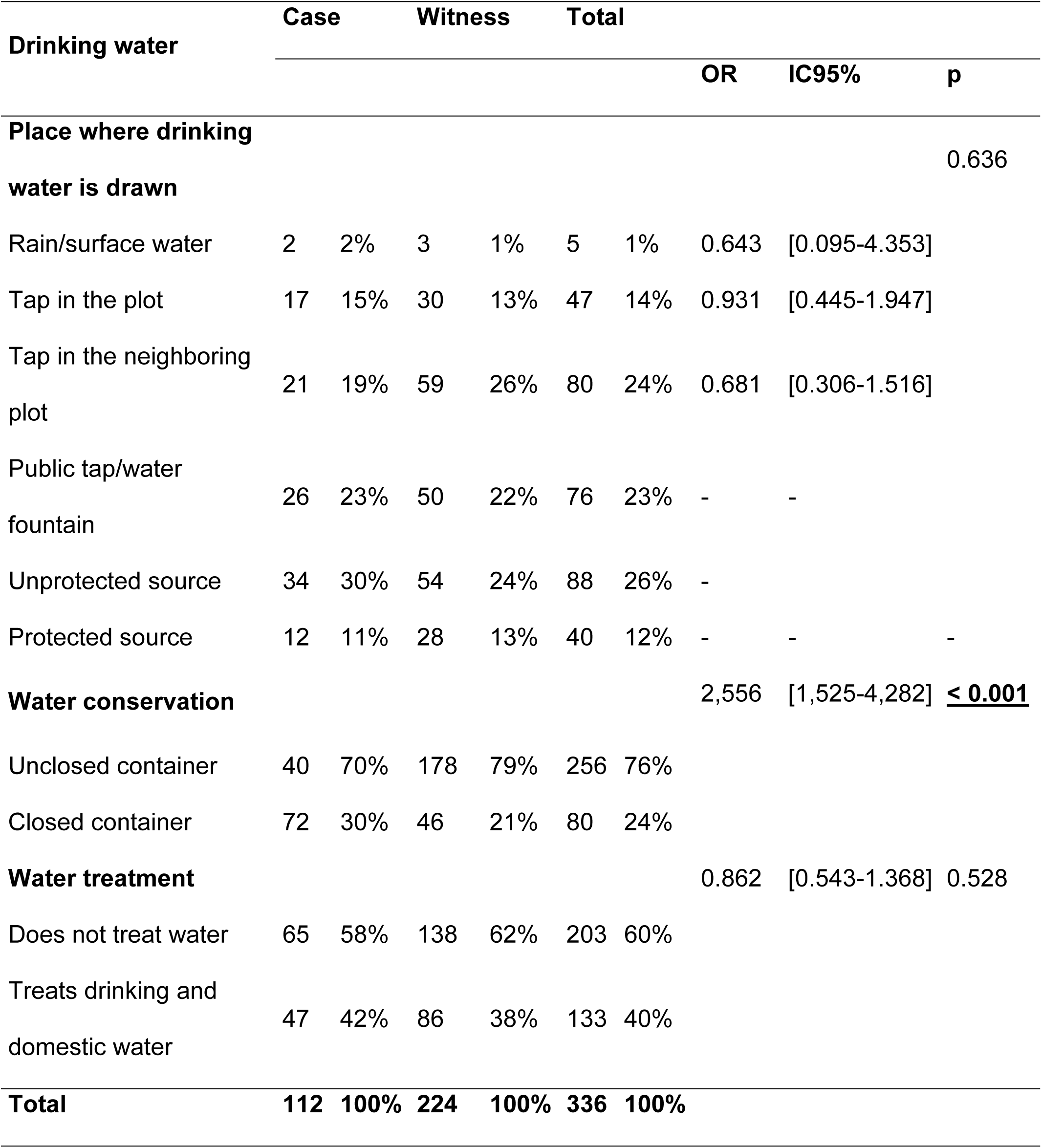
Location of drawing, storage and treatment of drinking water.

### III.4. The situation of water, sanitation and hygiene (WASH) in households

Table 3 shows that 38% of cholera patients do not take into account hygienic measures in food preservation against 18% who did not develop cholera (controls). Poor food preservation is a risk factor significantly associated with the cholera epidemic in the Kadutu HZ. OR = 2.678; 95% CI [1.607-4.464] p <0.001. The majority of cases (96%) do not wash their hands regularly before eating against 28% of controls. Failure to wash hands before eating is significantly associated with the cholera epidemic in the Kadutu HZ with OR = 8.190 95% CI [3.189-21.036] p <0.001. Nearly 2 out of 10 (21%) cases reported that lactating women do not wash their hands before breastfeeding their babies compared to 41% of controls. Non-washing of hands before breastfeeding is significantly associated with cholera outbreak in Kadutu HZ. OR = 3.321 95% CI [1.700-6.487] p <0.001. After toileting, 54% of cases do not wash their hands compared to 42% of controls. Non-washing of hands after toileting and non-washing of hands after contact with dirt are respectively significantly associated with cholera outbreak in Kadutu HZ. OR = 1.596 95% CI [1.011-2.519], p = 0.044 and OR = 6.929 95% CI [4.113-11.673] p < 0.001.

**Table 3:**
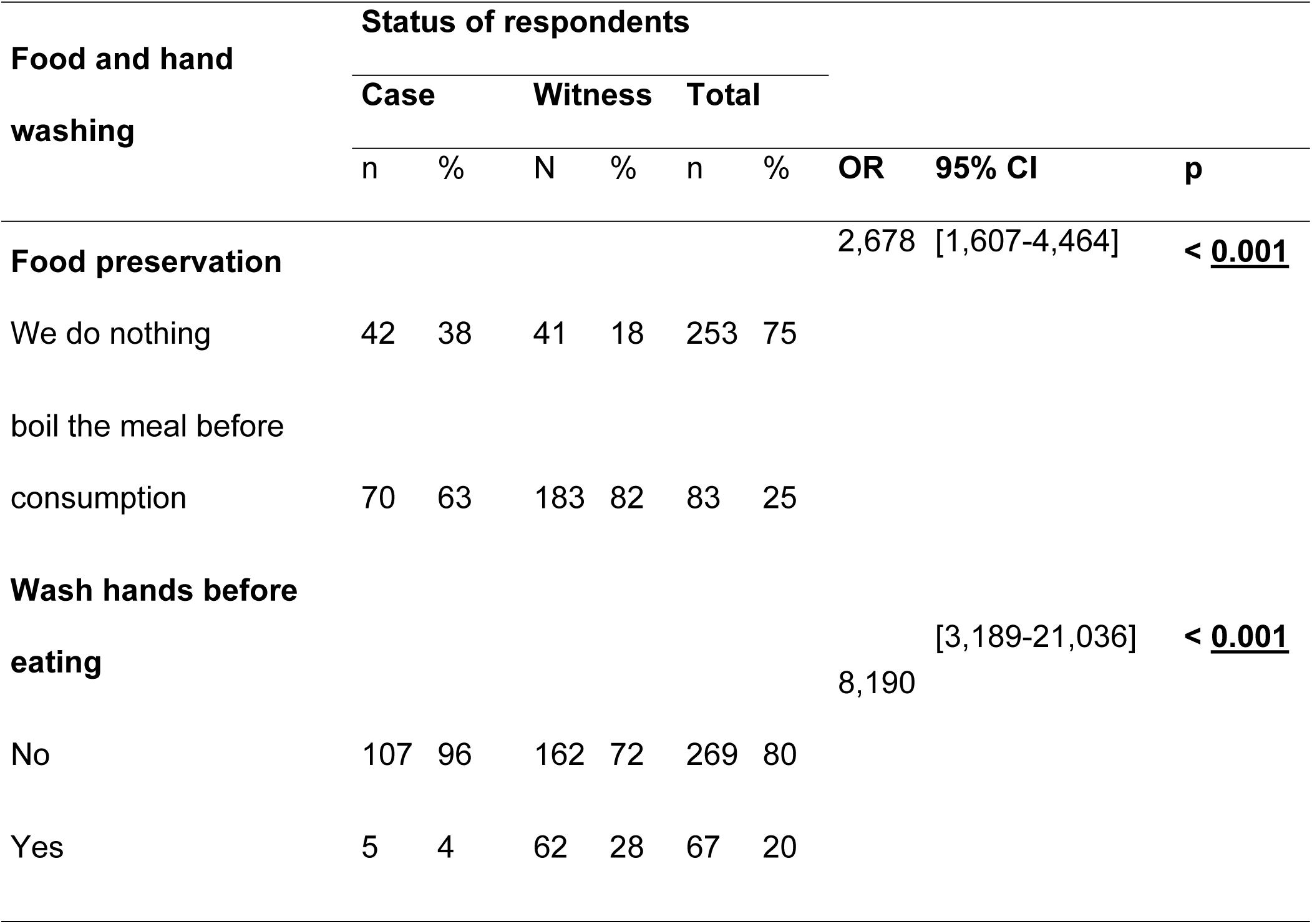
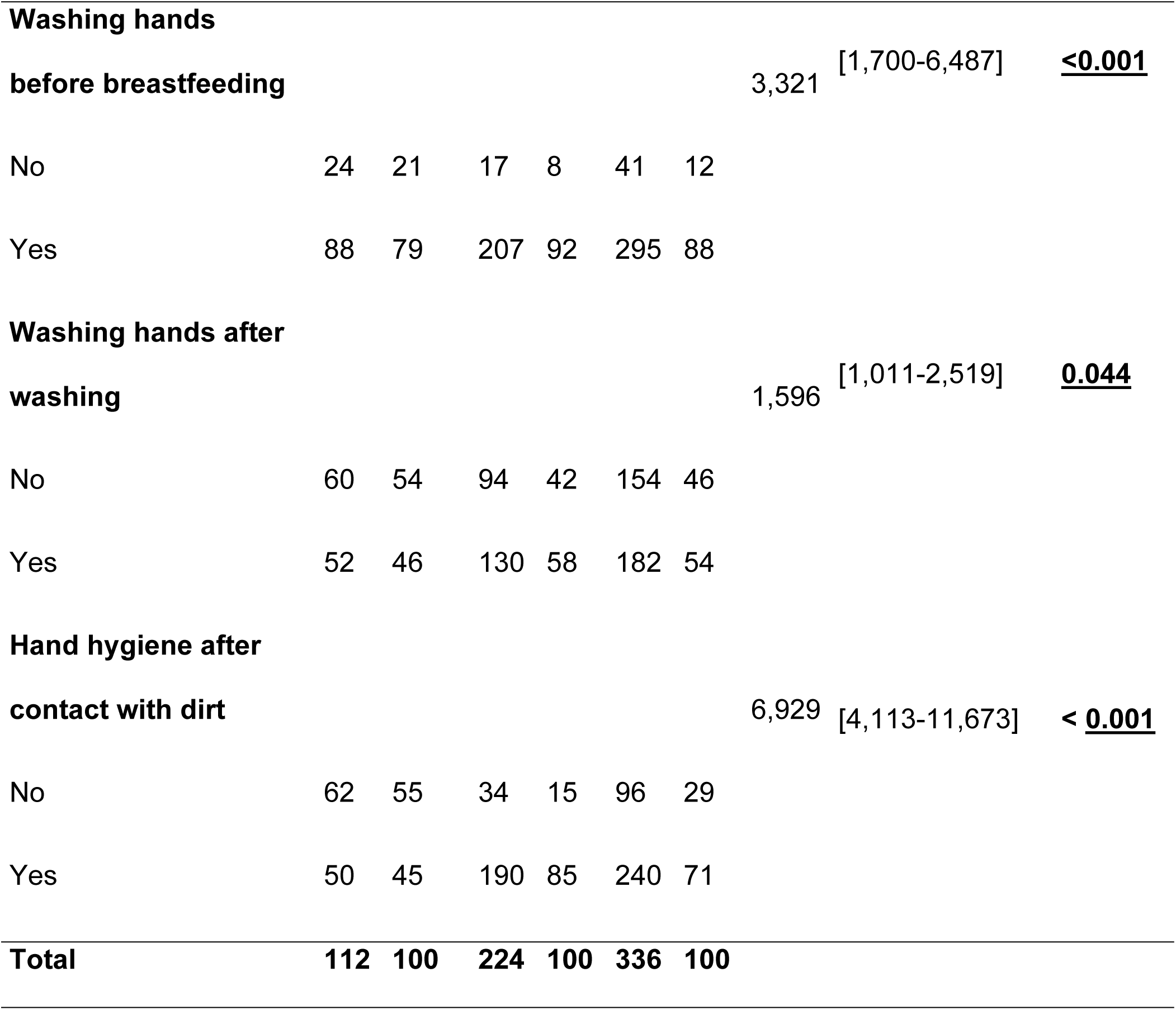
The situation of water, sanitation and hygiene (WASH) in households.

### III.5. Sanitation Conditions, Type of Toilet Flooring, and Climate Associated with Cholera in the Kadutu Health Zone

Table 4 shows that 30% of cases do not have toilets in the household compared to 17% of controls. The absence of toilets in the household is a factor that is associated with the cholera epidemic in the Kadutu HZ with OR = 2.203 95% CI [1.290-3.763] p = 0.003. However, 63% of cases and 50% of controls have unclean toilets. Unclean toilets in households are not a factor that can be associated with the cholera epidemic because OR = 1.004; 95% CI [0.372-2.710] p = 0.994. More cholera cases occurred during the rainy season, or 52% of cases. The cholera epidemic in the Kadutu HZ is independent of the season p = 0.334.

**Table 4:** Toilet location, floor type where the toilets are installed and climate associated with cholera in the Kadutu HZU.

| Sanitary facilities | Case |  | Witness |  | Total |  | OR | 95% CI | p |
| --- | --- | --- | --- | --- | --- | --- | --- | --- | --- |
| **Toilet in households** |  |  |  |  |  |  | 2,203 | [1,290-3,763] | **0.003** |
| No | 34 | 30% | 37 | 17% | 71 | 21% |  |  |  |
| Yes | 78 | 70% | 187 | 83% | 265 | 79% |  |  |  |
| **Dirty toilet** | **n=78** |  | **n=123** |  | **n=201** |  | 1,004 | [0.372-2.710] | 0.994 |
| No | 7 | 6% | 11 | 5% | 18 | 5% |  |  |  |
| Yes | 71 | 63% | 112 | 50% | 183 | 54% |  |  |  |
| **Type of soil where it is installed the toilet** |  |  |  |  |  |  |  |  |  |
| Clayey | 78 | 100% | 127 | 100% | 205 | 100% |  |  |  |
| **Epidemic season** |  |  |  |  |  |  | 0.882 | [0.560-1.390] | 0.588 |
| Rainy season | **58** | 52% | 123 | 55% | 181 | 54% |  |  |  |
| Dry season | 54 | 48% | 101 | 45% | 155 | 46% |  |  |  |
| **Total** | **112** | **100%** | **224** | **100%** | **336** | **100%** |  |  |  |

### III.6. Cultural and health factors of respondents in the Kadutu HZ

It appears from Table 5 that 85% of cases and 96% of controls confirm that the Cholera Treatment Center/Cholera Treatment Unit (CTC/CTU) is the facility responsible for managing cholera cases, while 15% of cases and 4% of controls indicate traditional healers as the location for cholera care. This culture is a factor significantly associated with the cholera epidemic in the Kadutu health zone where the OR = 3.829 95% CI [1.681- 8.674] and p = 0.001. Bad practice is observed in 79% of cases concerning the handling of cholera corpses before burial compared to 81% of witnesses. Sociocultural practices around a cholera corpse are not significantly associated with the epidemic in the Kadutu health zone where OR = 0.871; 95% CI [0.497-1.526], p = 0.678. Open defecation (OD) exposes the population to cholera 5.195 times in the Kadutu health zone where 63% of cases versus 25% of controls defecate outside the toilet. (OR = 5.195; 95% CI [3.185- 8.473]; p < 0.001).

**Table 5:**
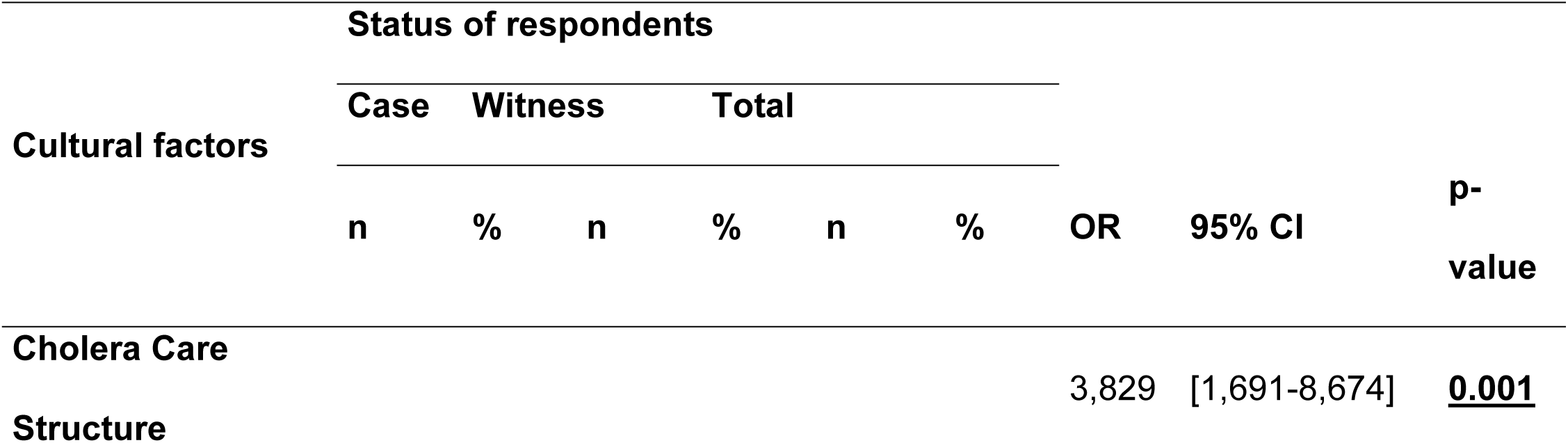
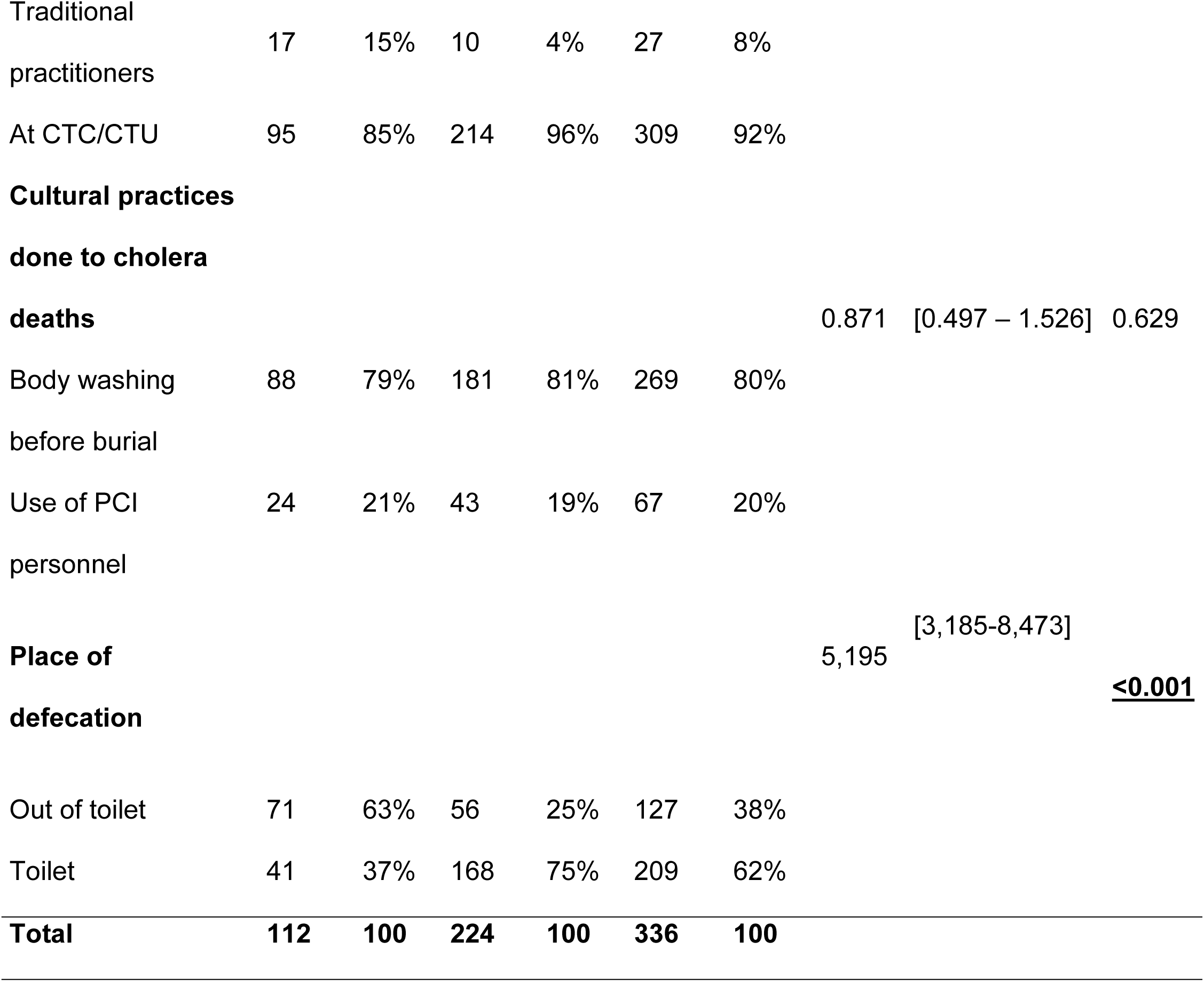
Cholera management, management of dead bodies and practices Defecation of people.

### III.7. Knowledge Analysis, Surveillance System and Cholera Vaccination Coverage in Kadutu Health Zone

Table 6 reveals that the modes of transmission of cholera are not well known by 11% of cases against 4% of controls. This is a factor significantly associated with the cholera epidemic in the HZK with OR = 2.568; 95% CI [1.074-6.143], p = 0.029. There is poor knowledge of the mode of prevention and treatment of cholera, 54 % of cases against 42% of controls, this ignorance is a risk factor significantly associated with the cholera epidemic in the HZK (OR = 1.596; 95% CI [1.011-2.519] p = 0.044). It turns out that only 2% of cases and 7% of controls were vaccinated against cholera. Non-vaccination is a factor significantly associated with the cholera epidemic in the HZK (OR = 4.231; 95% CI [0.955- 18.735] p = 0.040).

**Tableau 6.**
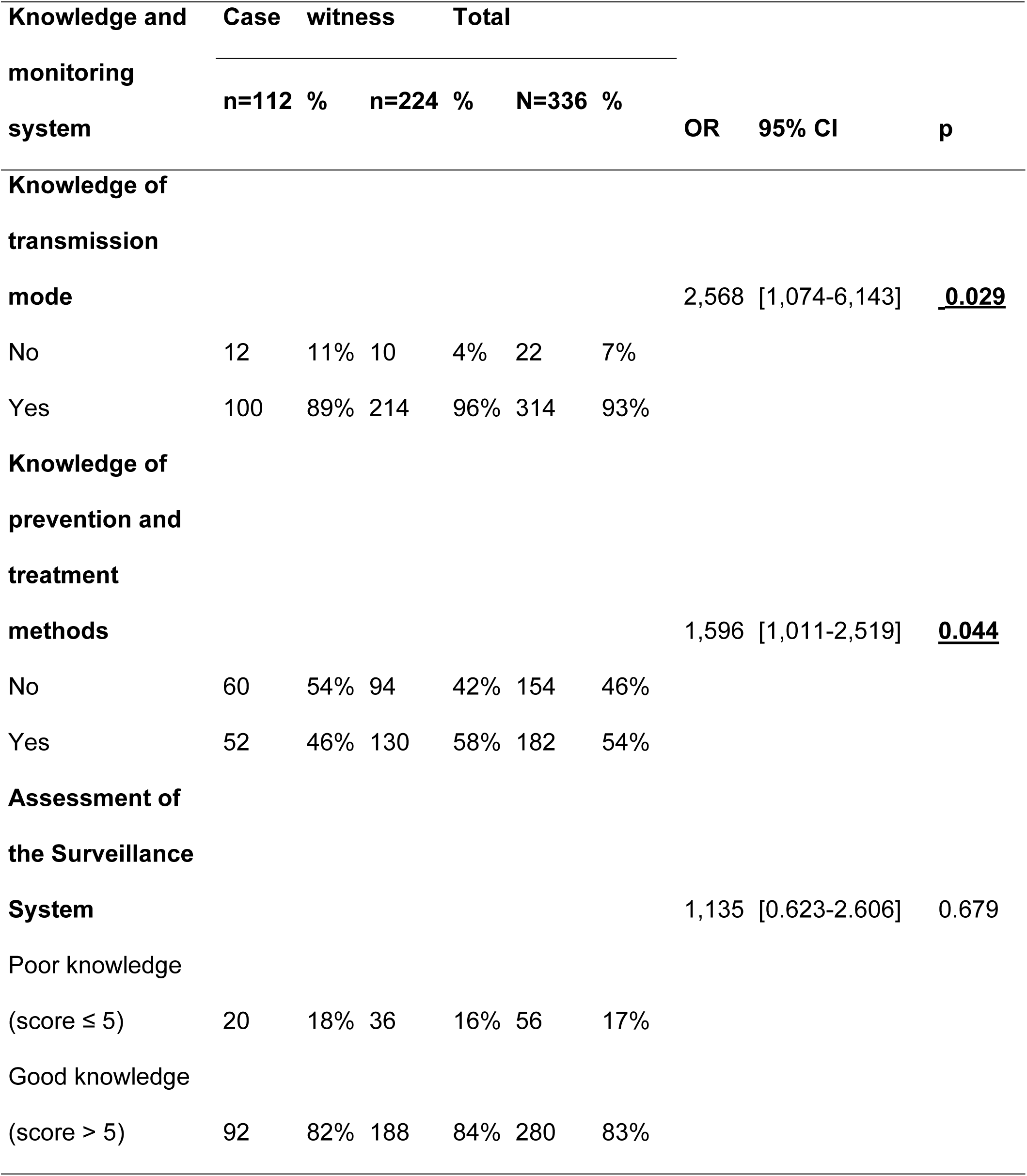
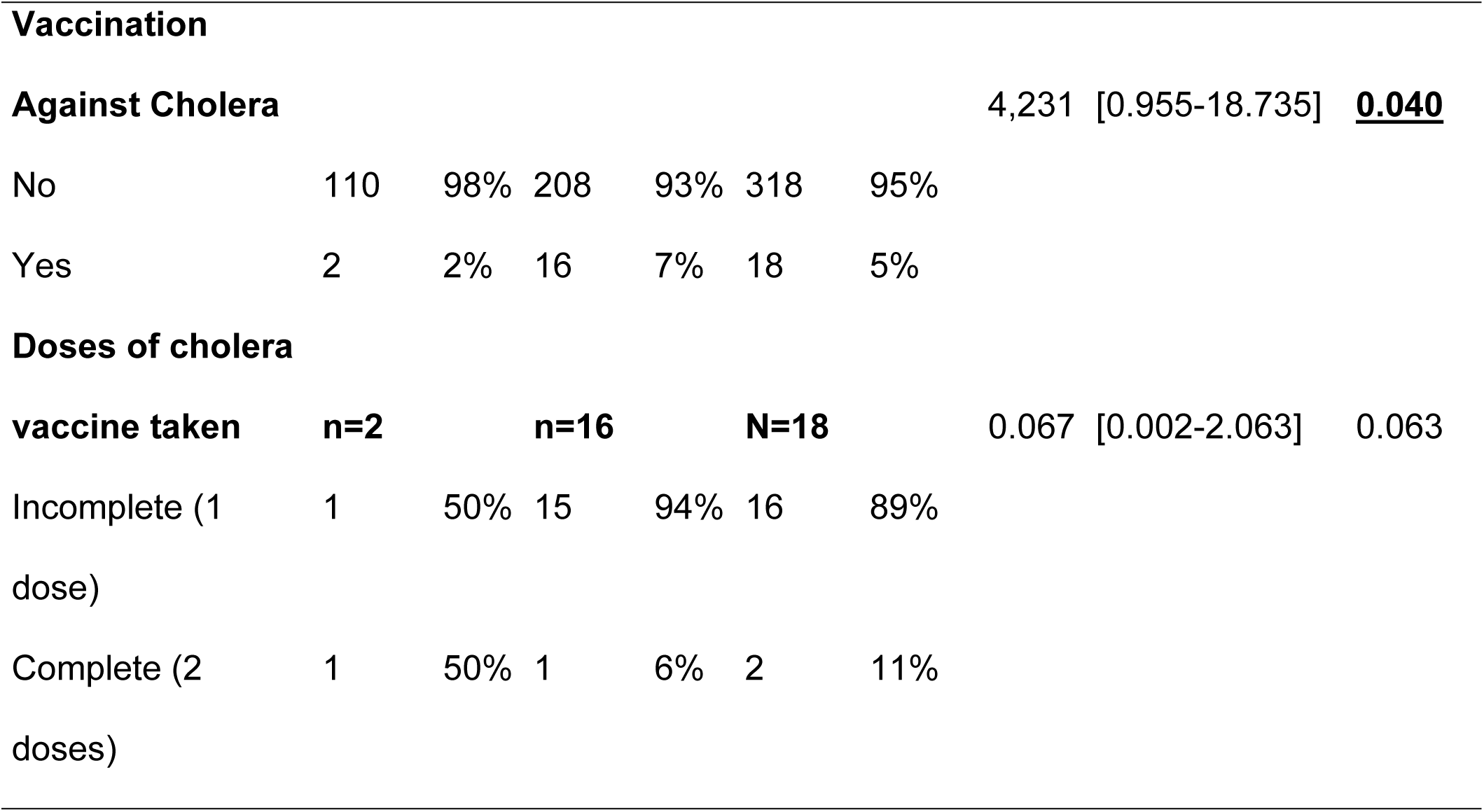
Knowledge Analysis, Surveillance System and Cholera Vaccination Coverage in Kadutu Health Zone.

### III.8. Factors associated with the persistence of cholera in the Kadutu HZ in 2023

Table 7 reveals, by bivariate analysis, fourteen factors that statistically explain the endemicity of cholera in the Kadutu HZ. These 14 factors are promiscuity; OR = 1.731; 95% CI [1.079 -2.777] p = 0.022]. Monthly income with an OR = 1.759; 95% CI [1.027-3.014] p = 0.039. Hand washing before eating OR = 8.190; 95% CI [3.189-21.036] p <0.001. Hand washing before breastfeeding the baby with an OR = 3.321 and its 95% CI of [1.700-6.487] p <0.001. Hand washing after bathing with an OR = 1.596 and its 95% CI of [1.011-2.519] p = 0.044. Hand hygiene after contact with dirt OR = 6.929 95% CI [4.113- 11.673] p < 0.001. Cholera PEC structure with an OR = 3.829 and its 95% CI of [1.681- 8.674] p = 0.001. Place of defecation; OR = 5.195; 95% CI [3.185-8.473] p < 0.001. Knowledge of the mode of transmission of cholera; OR = 2.568; 95% CI [1.074-6.143] p = 0.029. Knowledge of the mode of prevention and treatment of cholera with an OR = 1.596, the CI 95 [1.011-2.519] p = 0.044. Food preservation with an OR = 2.678 and its CI 95% ranging from [1.607-4.464] p < 0.001. The presence of toilets in the household with an OR = 2.203 and the CI 95% of [1.290-3.763] and p < 0.001. Vaccination against cholera OR = 4.231; 95% CI [0.955-18.735] p = 0.040 and finally the conservation of drinking water with an OR = 2.556 and the 95% CI [1.525-4.282] p = 0.046 and the p < 0.001.

**Tableau 7.**
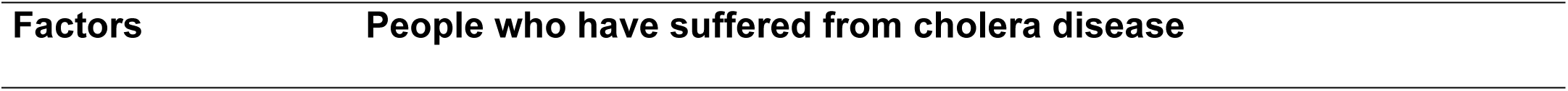
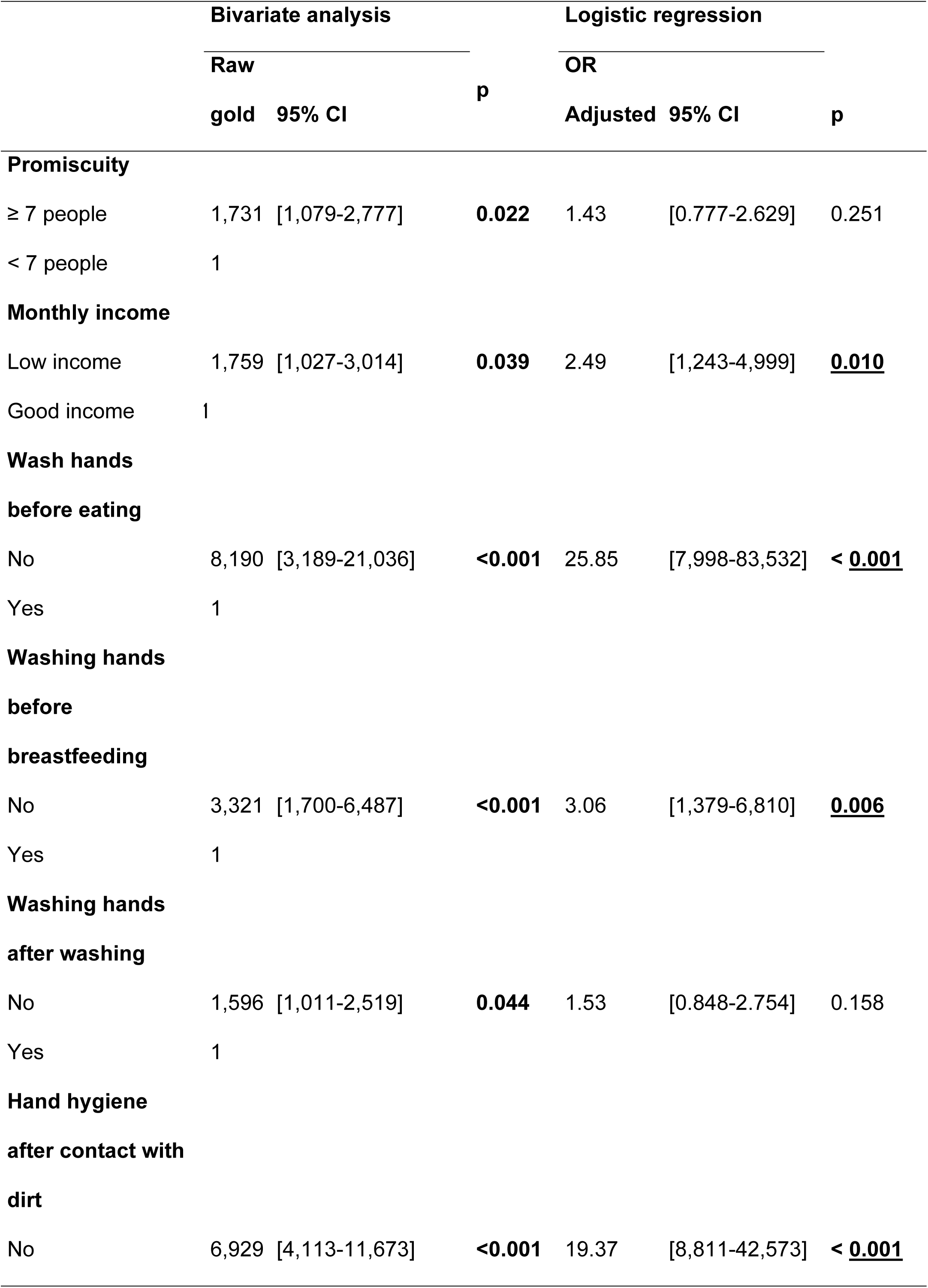
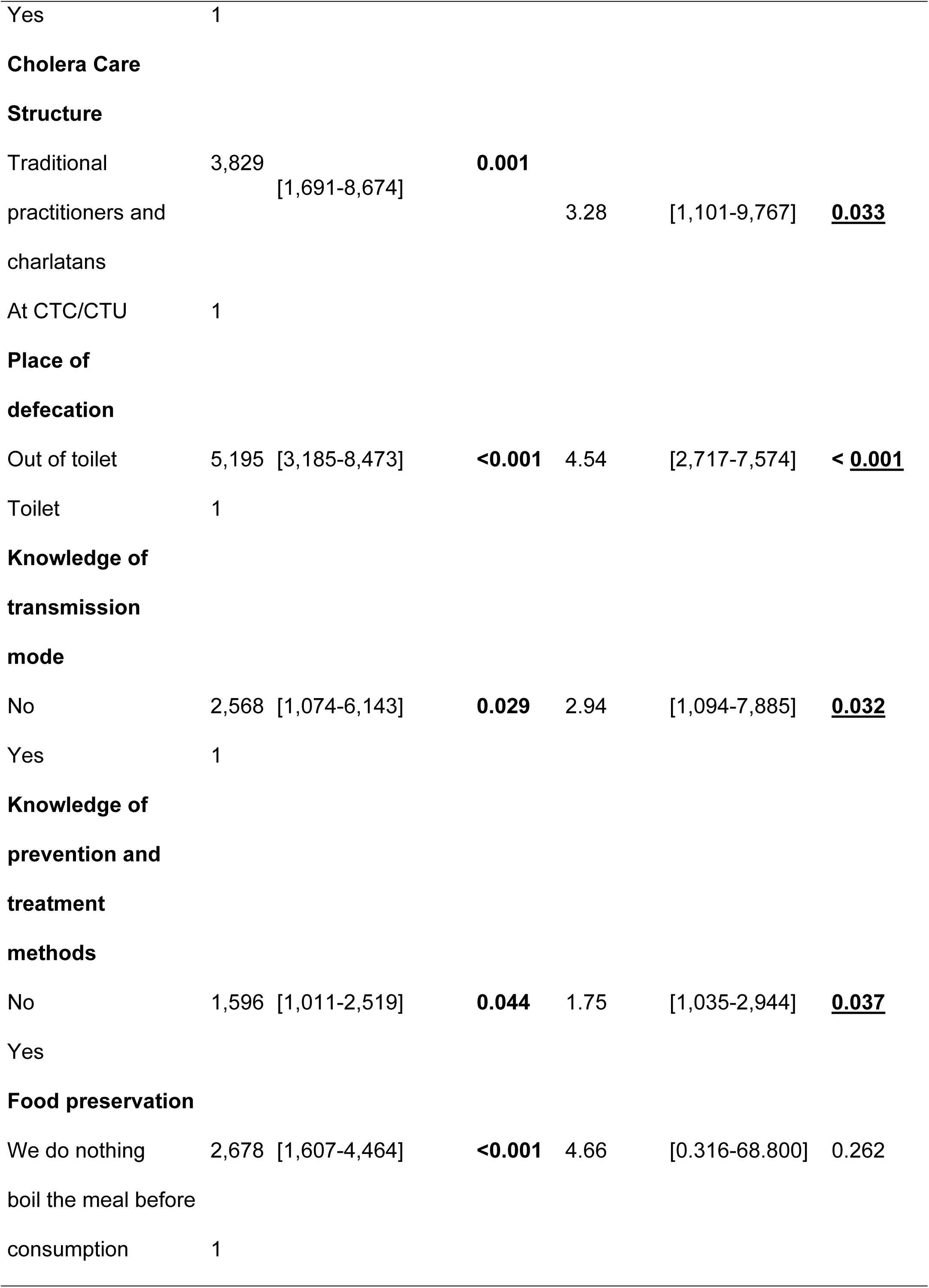
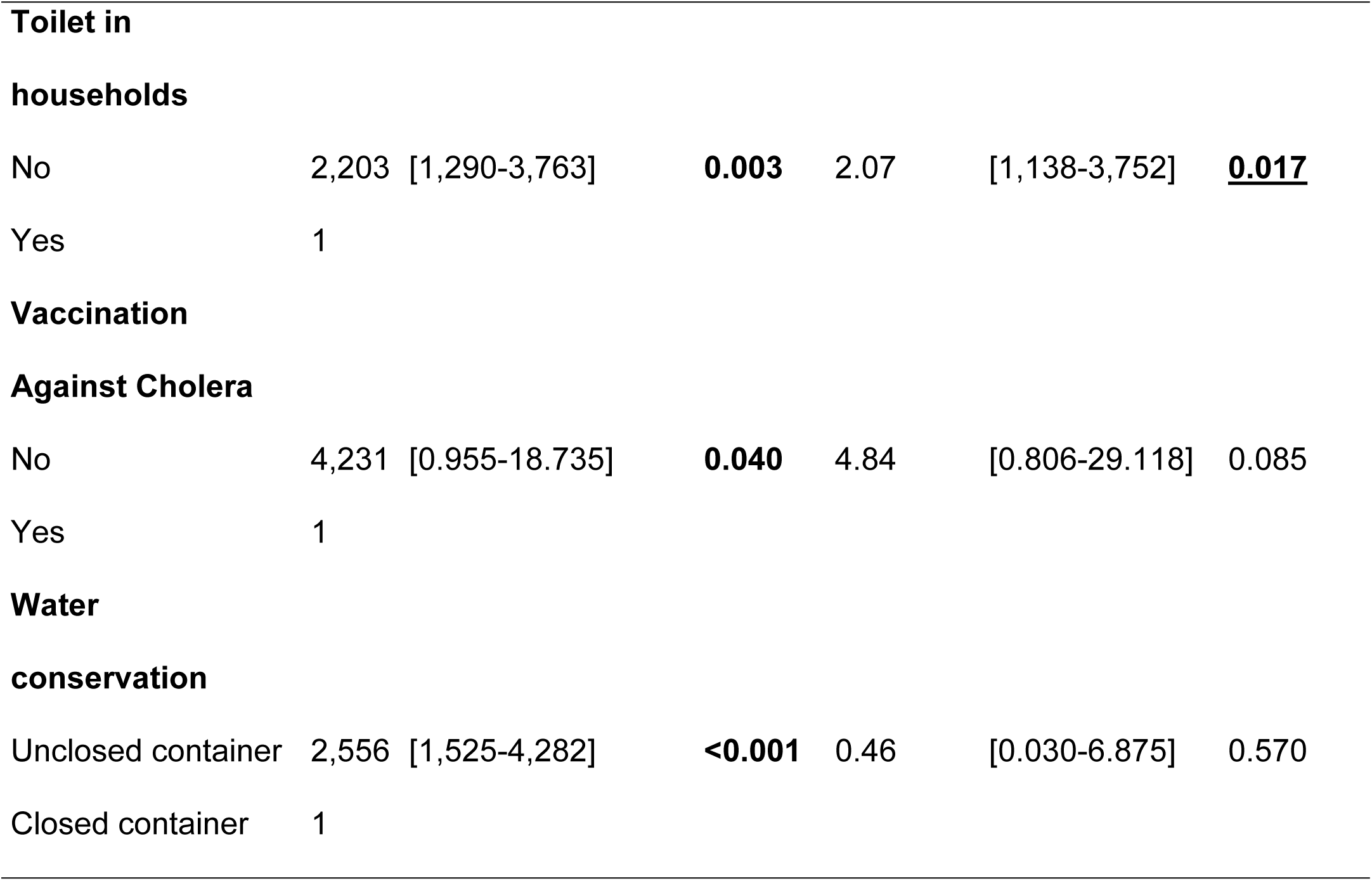
Promiscuity, monthly income, water, sanitation and hygiene situation in households in the Kadutu HZ associated with cholera.

However, logistic regression shows nine factors that are statistically significant. These factors are: Low monthly household income; OR = 2.49; 95% CI [1.243-4.999] p = 0.010. Failure to wash hands before eating OR = 25.85; 95% CI [7.998-83.532] p < 0.001. Failure to wash hands before breastfeeding the baby with an OR = 3.06 and its 95% CI of [1.379- 6.810] p = 0.006. Lack of hand hygiene after contact with dirt OR = 19.37 95% CI [8.811- 42.573] p < 0.001. The use of traditional practitioners and charlatans for the management (PEC) of cholera with an OR = 3.28 and its 95% CI of [1.101-9.767] p = 0.033. Defecation outside the toilet; OR = 4.54; 95% CI [2.717-7.574] p < 0.001. Lack of knowledge of the mode of transmission of cholera; OR = 2.94; 95% CI [1.094-7.885] p = 0.032. Lack of knowledge of the mode of prevention and treatment of cholera with an OR = 1.75, 95% CI of [1.035-2.944] and p = 0.037 and finally the lack of toilets in the household with an OR = 2.07 and 95% CI of [1.138-3.752] and p = 0.017. These factors predict cholera endemicity in the Kadutu HZ.

The number of people in the household, hand washing after washing, food preservation and cholera vaccination are not factors that explain the endemicity of cholera in the Kadutu HZ (p>0.05).

## IV. DISCUSSION

This study aims to identify the factors that can explain the occurrence of the cholera epidemic in the Kadutu health zone.

### Explanatory factors for the occurrence of the cholera epidemic in the Kadutu HZ

The result of the study shows that 42 % of cases and 29% of controls live with more than 7 people in the household. Overcrowding in households is not a factor that explains the endemicity of cholera in the Kadutu HZ with OR = 1.43 95% CI [0.777- 2.629], p = 0.25. Households with multi-storey houses with several rooms for its members are currently observed in the Kadutu HZ.

These results differ from the results of Muhumu P. et al., who found that there is a statistically significant difference between the occurrence of cholera and the size of households with an OR = 11.86 and its 95% CI [4.12-34.14] p = 0.0001 [9]. Several households host internally displaced persons. The displacement of the population is due to natural disasters and political instability in the region.

A household with low income is likely to be prone to cholera spread in Kadutu HZ with a 2.49 times risk of contracting cholera. OR = 2.49; 95% CI [1.243-4.999] p = 0.010. The poor are often exposed to unsanitary living conditions, favoring the spread of cholera. These results are similar to those obtained by Kibamba F. et al., in Uvira in 2019. Low-income households had a 0.45 times risk of contracting cholera due to poor waste management [10]. The socioeconomic context justifies this similarity in these two areas of the same province.

Building local capacity and investing in sustainable solutions are key to combating poverty and water-related diseases.

The results of this study did not show a significant difference in cholera and hand washing after toileting in households (OR = 1.53 and its 95% CI of [0.848-2.754] p = 0.158). However, it should be interpreted with caution, the latter should be guided by epidemiological knowledge and public health perspective. Hand washing after toileting may not be systematic in all households studied, which could make the results less significant, despite the theoretical recognition of its effectiveness in combating dirty hand diseases.

This result is opposed to that of Challa Jemal M et al. who found that not washing hands after washing exposes 3.25 times the risk of contracting cholera OR 3.04, 95% CI [1.58- 5.86] [11]. The result of Diner au G. et al. also found that washing hands with soap after washing is a protective factor for cholera OR: 0.04, 95% CI: [0.01, 0.25] [12]. Failure to take into account confounding factors, various modes of cholera transmission and environmental conditions unfavorable to *Vibrio cholerae* are among the factors that can explain these disagreements.

A more global, multifactorial approach can determine the link between cholera and handwashing practices.

In the present study, on the one hand, 85% of cases and 96% of witnesses confirmed that the CTC/CTU was a structure par excellence for the management (PEC) of cholera cases. However, on the other hand, 15% of cases and 4% of witnesses point to the structures of traditional practitioners as being the best place for the management of cholera.

The cholera PEC structure is an essential factor in explaining the occurrence of the cholera epidemic in the HZK with OR = 3.28 and its 95% CI of [1.101-9.767] p = 0.033. Cholera patients prefer to consult structures close to their households, these are private health care establishments (ESS), traditional practitioners’ offices. Others resort to self-medication for socioeconomic and cultural reasons. Pre-positioning a medical ambulance and raising awareness in the community about the use of CTC/CTU in case of diarrhea remains a priority in the Kadutu HZ.

Lack of hand hygiene after contact with dirt exposes its perpetrator to 19.37 times the risk of contracting cholera (OR = 19.37 95% CI [8.811-42.573] p <0.001). Dirty hands are a major vector of cholera transmission because they allow *Vibrio cholerae* to be transported and spread between the environment and the host. Cholera is mainly transmitted by the fecal-oral route.

These results are in disagreement with the findings of Hailu D et al. (2024) who found that hygiene was not associated with cholera OR=0.303 95% CI [0.273-0.333] p=0.36 [13]. However, the study setting context of southwest Ethiopia, with at least basic water and sanitation facilities, urban residents had better access to WASH facilities.

New public health and development measures, including improved water supplies and awareness raising, could impact hygiene practices.

The results of this study show that open defecation (OD) among the population exposes to a 4.54 times greater risk of being contaminated by cholera in the HZK (OR = 4.54; 95% CI [2.717-7.574] p <0.001). Excrement left in the open air, in the environment, contaminates the soil, rivers, the lake, domestic water sources and facilitates the spread of *Vibrio cholerae.* Flies, attracted by excrement, are vectors of bacteria and carry them to food. This phenomenon, combined with a lack of sanitation and appropriate hygiene practices, contributes to the transmission of cholera, particularly in vulnerable communities.

These results are also reported in other studies where the absence of hygienic latrines exposes household members 11 times more to being contaminated with cholera. OR = 11.65 with a 95% CI [7.73 - 17.63] with a p value < 0.05 [14].

The results of this study confirmed the empirical hypothesis that the culture of defecating outside the toilet contributes to cholera endemicity. It is essential to invest in sustainable sanitation infrastructure, such as public and private toilets, in order to limit OD.

The present study demonstrates that ignorance of cholera prevention and treatment methods is a risk factor statistically associated with cholera (OR = 1.596, CI 95 [1.011- 2.519] p = 0.044). The lack of sufficient awareness of hygiene practices and water resource management, leaving communities vulnerable to significant health risks, such as cholera.

This result is contrary to that of Nasr et al.2024 who found that there was no association between knowledge of preventive measures for cholera and cholera epidemic (OR = 0.9; 95% CI, [0.45-1.76]; p = 0.74) [15].

In addition, educational interventions on cholera can improve community knowledge and contribute to the prevention of cholera outbreaks.

Absence of toilets in the household was also identified as a risk factor for cholera in the HZK with an OR = 2.07 and 95% CI of [1.138-3.752] p = 0.017 [16]. *Vibrio Cholerae* carriers were more likely to defecate in the open, in the river, at the lake than controls.

In contrast to the results found in the Buea health district in Cameroon in 2015 (OR = 0.69, CI: 0.25–1.86, p = 0.490), cholera patients who defecated in bushes and rivers had the same odds of being infected with cholera as participants without the disease.

Without toilets, people are forced to defecate outdoors, which can contaminate the environment, including water, soil and food, increasing the risk of spreading cholera, a faecal-borne disease.

Non-vaccination against cholera is not a factor that explains the endemicity of cholera in the Kadutu HZ OR = 0.46 95% CI [0.030-6.875] p = 0.570. The recent cholera vaccination campaign in this area had encountered logistical constraints.

Cholera vaccine is a valuable tool in the fight against this disease in endemic areas. It helps reduce the number of cases and protect vulnerable populations in high-risk contexts [17].

The HZK, like many high-risk cholera regions, has logistical, political or financial obstacles that can limit access to the cholera vaccine. The vaccine, although effective in the short term, provides protection that generally lasts between 2 and 3 years. This requires regular boosters, especially in endemic areas such as the Kadutu HZ.

### IV.1. Strengths of the study

The unique strength of this study lies in the determination of the explanatory factors for the occurrence of the cholera epidemic in the Kadutu health zone in the city of Bukavu.

### IV.2. Limits of the study

Biological confirmation was not systematically carried out in all cholera patients, however compliance with the case definition was essential in order to correct the probable selection bias.

### IV.3. Conflicts of interest

The authors declare that they have no conflicts of interest.

### IV.4. Contribution of the authors

Bonhomme Kalimira Kachelewa initiated the study, developed the protocol and data collection tools. He ensured data collection, analysis, interpretation of results, and drafting and revision of the final manuscript. Denise Ngondo and Jean Nyandwe Kyloka participated in all stages of the study except data collection. Henry Mata, Jack Kokolomami, and Thiery Bobanga contributed to the final revision of the manuscript. All authors read the final version and gave their approval.

### IV.5. Acknowledgements

Our thanks go to the United States Agency for International Development (USAID) and the Regional Disease Surveillance Systems Strengthening Project, Phase 4 (REDISSE IV) for funding this study and work, to AFENET, to the authorities of the University of Kinshasa and the Kinshasa School of Public Health, to the Head of the Provincial Health Division of South Kivu, Dr. Gaston Maboko, to Dr. Aimé Alengo, Justin Bengehya, Dr Kasogo and to the PhD student Célestin Kyambikwa as well as to the teams of the Kadutu Health Zone for their administrative facilitation in the field. Finally, we extend our sincere thanks to Ms. Claudine Balindamwami for his encouragement and support.

## V. CONCLUSION AND RECOMMENDATIONS

### IV.1. CONCLUSION

The study on the explanatory factors of the cholera epidemic in the city of Bukavu, more particularly in the health zone of Kadutu, had the general objective of determining the factors which could explain the cholera epidemic in this Zone.

The study revealed nine factors explaining the endemic nature of cholera in the Kadutu HZ: Low monthly household income; not washing hands before eating; not washing hands before breastfeeding the baby; lack of hand hygiene after contact with dirt; recourse to traditional practitioners for PEC; defecation outside the toilet; ignorance of the mode of transmission, the mode of prevention and treatment of cholera and finally the lack of toilets in the household.

Promiscuity, hand washing after washing, food preservation and cholera vaccination did not emerge as factors significantly associated with the cholera epidemic in the Kadutu health zone.

Prevention of cholera epidemics in the Kadutu health zone remains multisectoral. It will take into account socio-demographic, economic, environmental, cultural and health factors.

### IV.2. RECOMMENDATIONS

In view of the results obtained, the following recommendations were made:

To the political and administrative authorities of the province :

- Strengthen local capacities, ensure access to health care and invest in sustainable solutions to poverty and water-related diseases.

At the Provincial Health Division (DPS) of South Kivu

- Support the HZ in implementing interventions aimed at improving access to drinking water, hygiene conditions and sanitation.

At the level of the Central Office of the Health Zone (BCHZ) Kadutu

- Strengthen community awareness on hand hygiene, non-exposure to open air and the importance of using CTC/CTU in case of diarrhea.

To the community:

- Adopt behaviors that reduce the risk of diarrheal diseases and cholera.

## Notes

### Competing Interest Statement

The authors have declared no competing interest.

### Funding Statement

The author(s) received no specific funding for this work.

### Author Declarations

The research protocol was submitted to the ethics committee of the Kinshasa School of Public Health (ESPK) and received its approval under N°. ESP/CE/189/2023. This study was conducted in strict compliance with the main ethical principles which are: respect for the person (autonomy and self-determination), and benevolence in justice.

